# Modeling COVID-19 latent prevalence to assess a public health intervention at a state and regional scale

**DOI:** 10.1101/2020.04.14.20063420

**Authors:** Philip J. Turk, Shih-Hsiung Chou, Marc A. Kowalkowski, Pooja P. Palmer, Jennifer S. Priem, Melanie D. Spencer, Yhenneko J. Taylor, Andrew D. McWilliams

## Abstract

**Background:** Emergence of COVID-19 caught the world off-guard and unprepared, initiating a global pandemic. In the absence of evidence, individual communities had to take timely action to reduce the rate of disease spread and avoid overburdening their healthcare systems. Although a few predictive models have been published to guide these decisions, most have not taken into account spatial differences and have included assumptions that do not match the local realities. Access to reliable information that is adapted to local context is critical for policymakers to make informed decisions during a rapidly evolving pandemic.

**Objective:** The goal of this study was to develop an adapted susceptible-infected-removed (SIR) model to predict the trajectory of the COVID-19 pandemic in North Carolina (NC) and the Charlotte metropolitan region and to incorporate the effect of a public health intervention to reduce disease spread, while accounting for unique regional features and imperfect detection.

**Methods:** Using the software package R, three SIR models were fit to infection prevalence data from the state and the greater Charlotte region and then rigorously compared. One of these models (SIR-Int) accounted for a stay-at-home intervention and imperfect detection of COVID-19 cases. We computed longitudinal total estimates of the susceptible, infected, and removed compartments of both populations, along with other pandemic characteristics (e.g., basic reproduction number).

**Results:** Prior to March 26, disease spread was rapid at the pandemic onset with the Charlotte region doubling time of 2.56 days (95% CI: (2.11, 3.25)) and in NC 2.94 days (95% CI: (2.33, 4.00)). Subsequently, disease spread significantly slowed with doubling times increased in the Charlotte region to 4.70 days (95% CI: (3.77, 6.22)) and in NC to 4.01 days (95% CI: (3.43, 4.83)). Reflecting spatial differences, this deceleration favored the greater Charlotte region compared to NC as a whole. A comparison of the efficacy of intervention, defined as 1 - the hazard ratio of infection, gave 0.25 for NC and 0.43 for the Charlotte region. Also, early in the pandemic, the initial basic SIR model had good fit to the data; however, as the pandemic and local conditions evolved, the SIR-Int model emerged as the model with better fit.

**Conclusions:** Using local data and continuous attention to model adaptation, our findings have enabled policymakers, public health officials and health systems to proactively plan capacity and evaluate the impact of a public health intervention. Our SIR-Int model for estimated latent prevalence was reasonably flexible, highly accurate, and demonstrated the efficacy of a stay-at-home order at both the state and regional level. Our results highlight the importance of incorporating local context into pandemic forecast modeling, as well as the need to remain vigilant and informed by the data as we enter into a critical period of the outbreak.

## Introduction

In December 2019, a novel coronavirus emerged in Wuhan, Hubei province, China [1]. The pathogen causes a respiratory illness, now known as the coronavirus disease 2019 (COVID-19) [2, 3]. From its original epicenter in Wuhan, the virus spread rapidly within 30 days to other parts of mainland China and also exported to other countries [4, 5, 6, 7, 8]. As of April 10, 2020, 210 countries and territories have reported 1,673,423 confirmed cases of COVID-19 and 101,526 deaths [9]. Due to the spread across multiple countries and the large number of people impacted, on March 11 the World Health Organization recognized the novel severe acute respiratory syndrome coronavirus 2 (SARS-CoV-2) as a pandemic that poses a major global public health threat [10, 11].

While the effects of the COVID-19 pandemic are experienced globally, many key health policy decisions designed to reduce transmissions are determined at national and regional levels. These critical policy decisions must be implemented quickly and evaluated continuously so they can be adapted to the local context, recognizing the clear effect that geography, community context, density and social determinants of health have on COVID-19 outcomes. In North Carolina (NC), the first COVID-19 case was reported on March 2, 2020, and cases increased to 3,963 total confirmed cases as of April 10 [12]. To slow the rapidly increasing transmission rate, within a few weeks after the 1st case was detected, NC state officials promoted social distancing strategies (i.e., deliberately increasing physical space), banned large social gatherings, and closed public schools and universities. Subsequently, a stay-at-home order, which only allows for essential travel outside the home, was issued in the southwestern part of the state by Mecklenburg County effective at 8 am March 26, lasting through April 16 (since extended to April 29), while a statewide stay-at-home order was issued effective at 5 pm March 30, lasting to April 29.

Because the COVID-19 landscape evolves rapidly due to the confluence of locally relevant factors, appropriate modeling using timely infection prevalence to drive decision making around containment, treatment, and resource planning is critical. Forecasting models are used to generate early warnings to identify how a pandemic might evolve. During the early stages of the COVID-19 pandemic, forecasting was frequently applied to predict national and international infection transmission trends [13, 14]. Local communities and health systems turned to these national and international models for their own planning; however, the generalizability of such models to the local situation is limited and ignores important community-level population characteristics and transmission dynamics [3, 15, 16, 17]. An objective of this study was to understand how spatial differences impact model results and their interpretation.

In response to the need for actionable data insights in our community and health system, investigators from the Atrium Health Center for Outcomes Research and Evaluation (CORE) developed a series of COVID-19 forecasting models, which were used to guide Atrium Health’s initial proactive response to ensure sufficient capacity to treat the expected surge in patient care demands. In this study, we present an initial Susceptible-Infected-Removed (SIR) epidemic model and its evolution to the Susceptible-Infected-Removed-Social Distancing-Detection Rate (SIR-Int) model. Here we describe and compare these models, the spatial differences in a pandemic, the significance of observed cases versus actual prevalence in the setting of rapidly evolving testing strategies, the current epidemiological trends and the potential effects of non-pharmaceutical interventions applied locally (e.g., social distancing).

## Methods

The observed cumulative case and death counts were obtained daily at noon starting March 2, 2020, when the first COVID-19 case was reported, from the North Carolina Department of Health and Human Services website for all 100 counties [12]. Data collection for this manuscript ended on April 7, just prior to submission. In order to accurately estimate the actual latent prevalence at time t, the cumulative case counts were adjusted for imperfect detection by dividing them by 0.14. While estimates of detection probability for coronavirus, also known as the ascertainment rate, vary in the literature, ours is in line with those reported [18, 19, 20, 21, 22]. Modeling only the observed prevalence will give an inaccurate timeline of pandemic behavior. Next, cumulative deaths were subtracted from adjusted cumulative cases. We also adjusted cumulative cases for recoveries by removing cases after 20 days, the estimated median duration of viral shedding from illness onset [23]. Daily incremental incidence was obtained by subtracting the estimated latent prevalence at time *t* − 1 from that at time t. Crucially, in our research, we model estimated latent prevalence as constructed here, not observed prevalence. For the sake of brevity moving forward, we use the terms ‘latent prevalence’ and ‘prevalence’ interchangeably.

In addition to North Carolina, interest also lay in the subpopulation served by Atrium Health’s greater Charlotte market. For convenience, we make use of a designation of a group of counties that constitute the greater Charlotte area used by the North Carolina Department of Health and Human Services. Specifically, the US Centers for Disease Control and Prevention’s Cities Readiness Initiative (CRI) is a federally funded program designed to enhance preparedness in the nation’s largest population centers in order to rapidly and effectively respond to large public health emergencies (e.g., act of bioterrorism). This also allowed us to model on a region that harmonized with the state’s approach to disaster planning in case statewide coordination of resources would be required. Within NC, 11 counties are grouped into a CRI region that includes Anson, Cabarrus, Catawba, Cleveland, Gaston, Iredell, Lincoln, Mecklenburg, Rowan, Stanly, and Union. Collectively, we henceforth refer to these counties as ‘the CRI’ (Figure 1). Because the CRI closely mirrors the large area served by Atrium Health’s greater Charlotte market, we used this population base for our local modeling efforts. The CRI includes over 2.5 million residents (24% of the NC population) and ranges from rural settings like Anson County to Mecklenburg County, which contains NC’s largest city, Charlotte. To understand how spatial differences impact model results and their interpretation, we compared the CRI to NC throughout the early phases of this pandemic.

**Figure 1:**
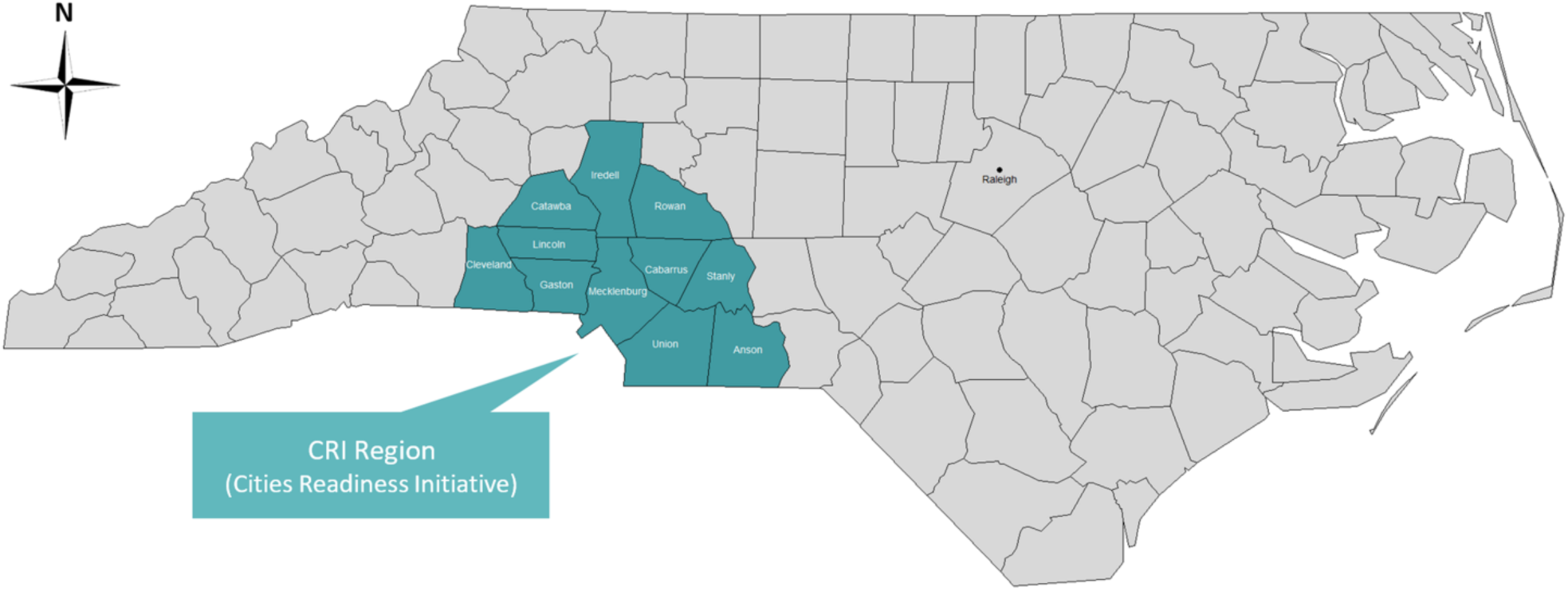
Map of North Carolina showing the CRI region.

We introduce the SIR deterministic compartmental model originally described by Kermack and McKendrick [24] and depict it in Figure 2.

**Figure 2:**
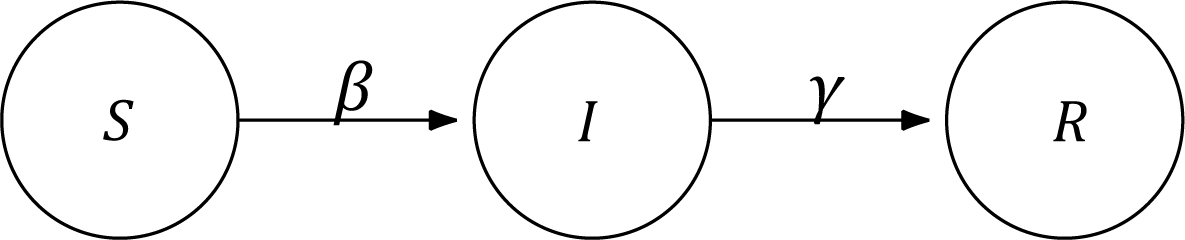
SIR model diagram showing compartments and flow.

*S* is the number of individuals that are <u>susceptible</u> to infection in the population, *I* is the number of individuals that are <u>infected</u>, and *R* is the number of individuals that are <u>removed</u> from the population via recovery and subsequent immunity or death from infection. This mutually exclusive and exhaustive partition is such that *S + I + R = N*, where *N* is the closed population size. We further assume all uninfected individuals are susceptible to infection. The transition flow is described by the arrows in the figure labeled with two rates. The parameter *β* is the infection rate and can be further decomposed as the product of the probability of transmission per contact and the rate of contact per person per unit time. *γ* is the removal rate.

More formally, the SIR model is a system of three ordinary differential equations (ODEs) involving two unknown parameters:

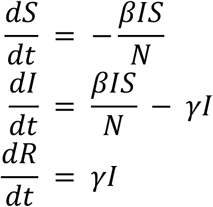

Note that all of *S, I*, and *R* and their derivatives are functions of time *t* (e.g., *S* = *S*(*t*)), although we do not denote this notationally here. By how the model is constructed, the first equation in the system returns a number less than or equal to zero, the second equation returns any real number, and the third equation returns a number greater than or equal to zero.

All data analysis was done using R statistical software, version 3.6.2. As described in Churches [25], we used the ode() default solver from the desolve package to solve the system of ODEs defining the SIR model. Next, we used a quasi-Newton method with constraints to find the optimal values for *β* and *γ* on (0, 1) by minimizing the square root of the sum of the squared differences between I, which is our prevalence, and its prediction 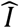 over all time *t* [26]. In order to establish initial conditions for model fitting, we estimate the population size of NC and the CRI to be 10,488,084 and 2,544,041, respectively, using information taken from census estimates. After obtaining the estimates 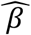 and 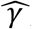, to help assess model goodness-of-fit, we define the following statistic:

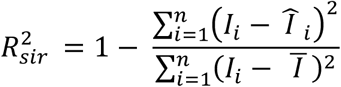

where time is indexed from *i =*1*,…,n*, and *n* is the number of prevalences in the sample. Note that *Ī* is the average of the *I_i_*’s.

In order to compare different scenarios, for both NC and the CRI, we define a SIR model (SIR-Pre) fit to the data from the time of the outbreak until the time of the March 26 Mecklenburg County stay-at-home order. Since Mecklenburg County is the state’s second largest county, this could have a strong effect on the pandemic trajectory, both in the CRI and the state; therefore, we have used this date to delineate the date of the significant public health intervention. We further define a SIR model (SIR-Post) fit to the data from the time of the outbreak until the end of data collection.

Given the major public health intervention implemented on March 26, we modify the SIR model for both the CRI and NC to accommodate this (denoted SIR-Int). SIR models with interventions can be simulated using the EpiModel package. This package provides tools for building, simulating, and analyzing several classes of models for the population dynamics of infectious disease transmission in epidemics. These include not only deterministic compartmental models, but stochastic individual contact models and network models. We first fit the SIR model as before to the data up until March 26, and extracted the estimates of *β* and *γ*. After March 26, we retained the removal rate, but modified the infection rate. First, we set the pre-intervention probability of transmission equal to 0.015, which is consistent with other viral infectious diseases like SARS and AIDS [27, 28]. We then set the rate of contact so that the probability of transmission multiplied by the rate of contact equaled 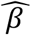. To simulate the observed intervention, using the default RK4 ODE solver, we affected the probability of transmission by iteratively decreasing the hazard ratio of infection given exposure to the intervention (step size of 0.0001) compared to no exposure, until the fitted infection curve yielded a maximum 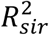.

For exploratory data analysis, we generated time plots for prevalence, incidence, and both daily and cumulative deaths. The basic reproduction number *R*_0_ is the average number of secondary cases of disease caused by a single infected individual over his or her infectious period in a population where all individuals are susceptible to infection. To estimate *R*_0_, we compute:

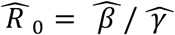

where 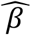 and 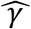 are estimates taken from the model fit. Since the SIR model is fully parameterized by *β* and *γ*, we also obtain predictions 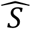 and 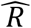 over all time *t*. The percentage of infected at peak prevalence was computed by dividing the maximum 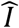 by the population size *N*, while the final percentage of infected was computed as the limit 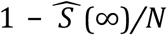. To estimate doubling time and compute a 95% confidence interval, we modeled incidence growth by fitting a loglinear model as a function of time *t* using the incidence package.

## Results

Figure 3 shows time plots of prevalence, cumulative deaths, incidence, and daily deaths for NC from the start of the outbreak on March 2 up to and including April 7. The first death was recorded in NC on March 24.

**Figure 3:**
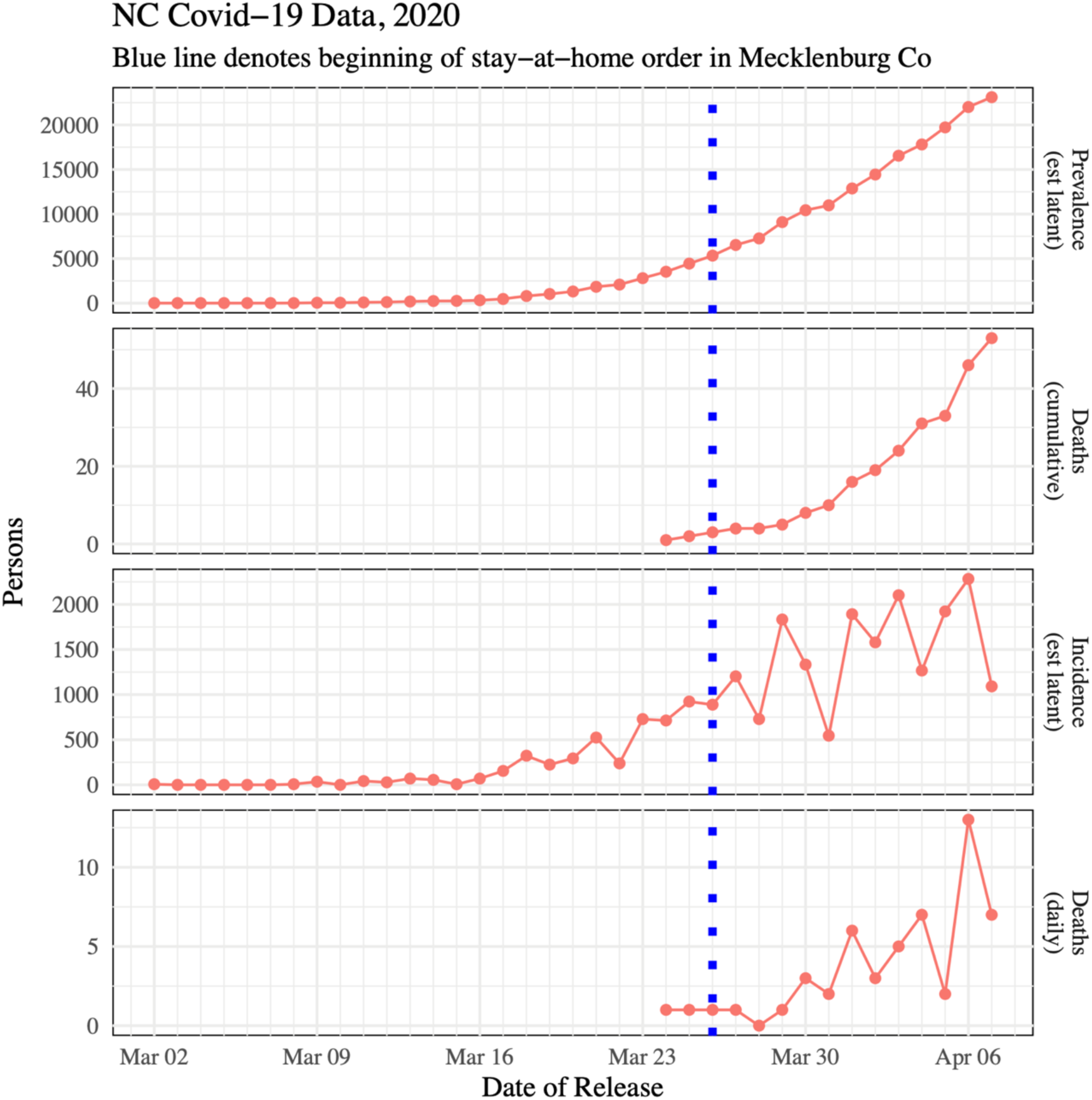
Time plots for NC.

Figure 4 shows time plots of prevalence, cumulative deaths, incidence, and daily deaths for the CRI from the start of the outbreak on March 11 up to and including April 7. The first death was recorded in the CRI on March 25.

**Figure 4:**
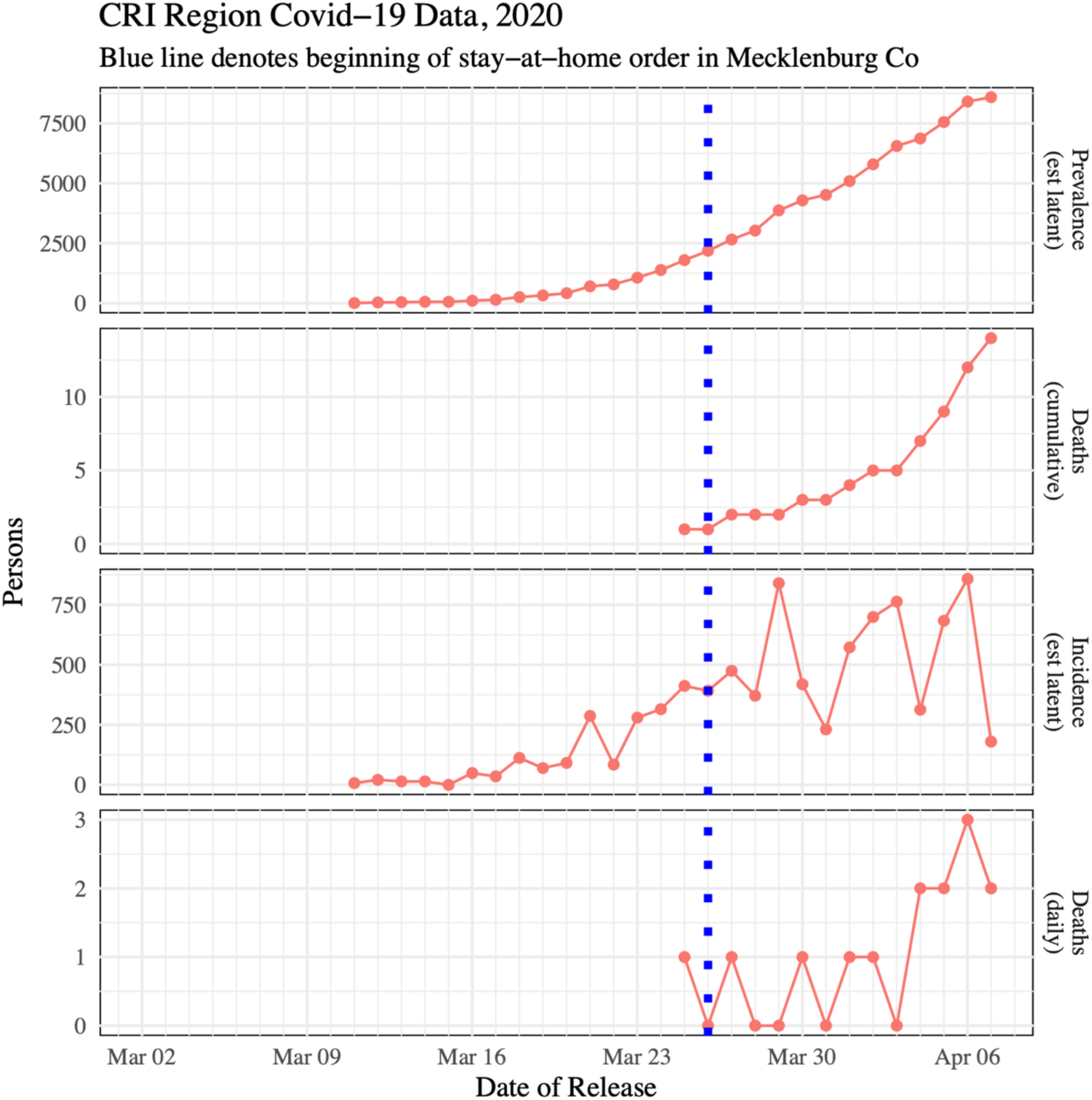
Time plots for the CRI.

Notably, the prevalence and cumulative death curves for both figures look exponential. While both incidence curves are increasing, the incidence curves become volatile after the stay-at-home order went into effect. Prior to March 26, doubling time was estimated to be 2.56 days in the CRI (95% CI: (2.11, 3.25)) and 2.94 days in NC (95% CI: (2.33, 4.00)). Once data after March 26 are included, the doubling times increased and were estimated to be 4.70 days in the CRI (95% CI: (3.77, 6.22)) and 4.01 days in NC (95% CI: (3.43, 4.83)).

Tables 1 and 2 gives a synopsis of the model fits for each location and model type. The estimated *R*_0_ of 2.36 for the CRI prior to March 26 is more typical of the range of *R*_0_ values given in the literature for COVID-19, while the value of 1.79 for NC is substantially lower [29, 30]. After the intervention, the estimated Ro values for both locations drop to a similar value, although this result is affected by reduced model fit. A comparison of the efficacy of intervention, defined as 1 - the hazard ratio of infection, gives 0.25 for NC and 0.43 for the CRI. Using these hazard ratios to compute estimates of *R*_0_ from March 26 onward (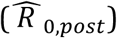), we derive 1.34 and 1.33 for NC and the CRI, respectively. This suggests the COVID-19 outbreak is rapidly decelerating in NC and the CRI after the aggressive public health intervention.

**Table 1:** Summary table of model fit for SIR-Pre and SIR-Post models in NC and the CRI.

| Location | Model | $\widehat{\beta}$ | $\widehat{\gamma}$ | $\widehat{R}_0$ | $R^2_{sir}$ |
| --- | --- | --- | --- | --- | --- |
| NC | SIR-Pre | 0.6415 | 0.3585 | 1.79 | 0.99 |
| NC | SIR-Post | 0.6165 | 0.3835 | 1.61 | 0.84 |
| CRI | SIR-Pre | 0.7020 | 0.2980 | 2.36 | 0.94 |
| CRI | SIR-Post | 0.6381 | 0.3619 | 1.76 | 0.65 |

**Table 2:** Summary table of model fit for SIR-Int model in NC and the CRI.

| Location | Hazard Ratio | $R^2_{sir}$ | $\widehat{R}_{0,post}$ |
| --- | --- | --- | --- |
| NC | 0.75 | 0.99 | 1.34 |
| CRI | 0.57 | 0.99 | 1.33 |

Figures 5 and 6 show plots of the three fitted models’ infection curves for NC and the CRI, respectively, out to April 7. The behavior in the two plots is the same. The SIR-Post model clearly demonstrates a lack-of-fit to the data. For the SIR-Int model, we note the hinge point induces a change of behavior from March 26 onward. The dotted orange line represents SIR-Pre forecast projections from March 26 onward. Note they are much larger than the actual data.

**Figure 5:**
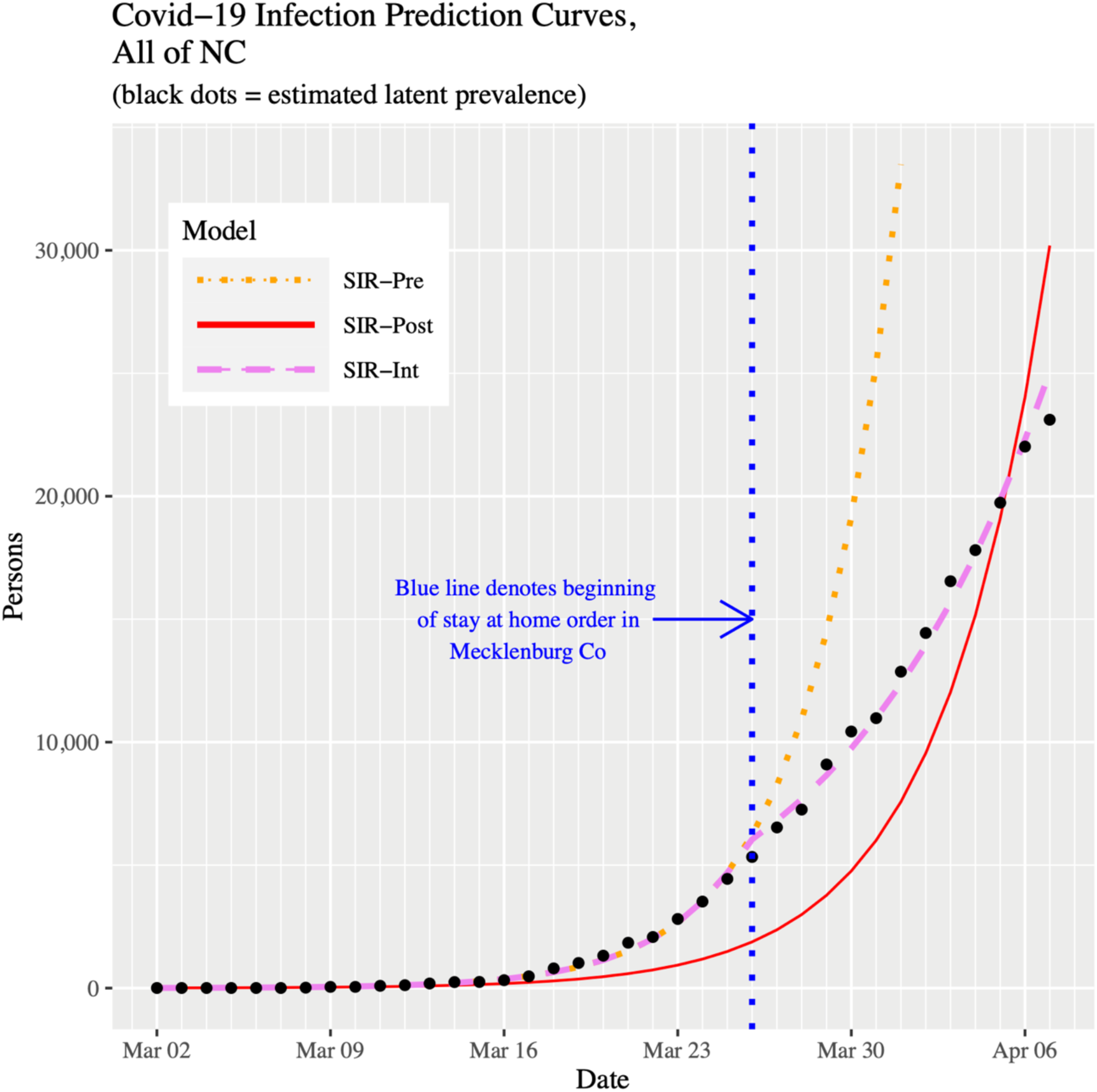
Infection prevalence prediction curves for NC up to April 7, 2020.

**Figure 6:**
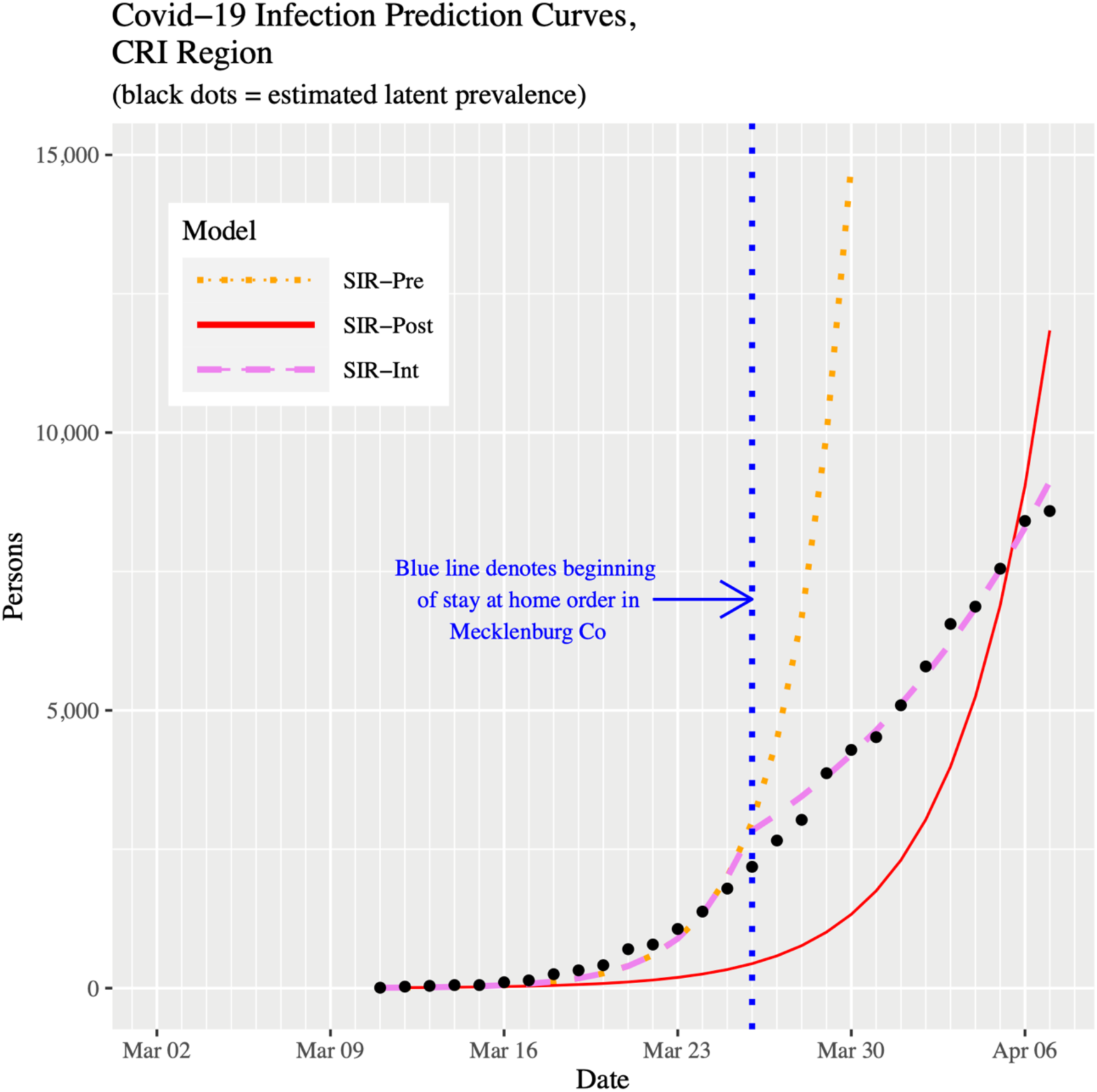
Infection prevalence prediction curves for the CRI up to April 7, 2020.

Figures 7 and 8 show plots of the three fitted models’ infection curves for NC and the CRI, respectively, projected out to the beginning of August. In both plots, we see the dramatic effect of the public health intervention; that is, the so-called “flattening of the curve”. There are two important differences to note between NC and the CRI region. First, the CRI visibly shows relatively more flattening. This effect can be best observed in Table 3 in the Peak % Infected and Final % Infected columns. Moving from the Pre to Post to Int models within a location, the drop in percentage infected is more pronounced in the CRI. In fact, for the SIR-Int model, the percentages are virtually the same for both locations; that is, the CRI has “slowed down” to the state as a whole. Second, the date of peak prevalence was initially 8 days earlier for the CRI compared to NC. However, using the current SIR-Int model, although both locations showed their infection curves shifting forward in time, the date of peak prevalence is now 3 days later in the CRI (Table 3). To put this into context, for NC, the time duration from the start of the outbreak to the peak prevalence has gone from 49 days to 70 days (43% increase). However, for the CRI, the time duration from the start of the outbreak to the peak prevalence has gone from 32 days to 64 days (100% increase).

**Figure 7:**
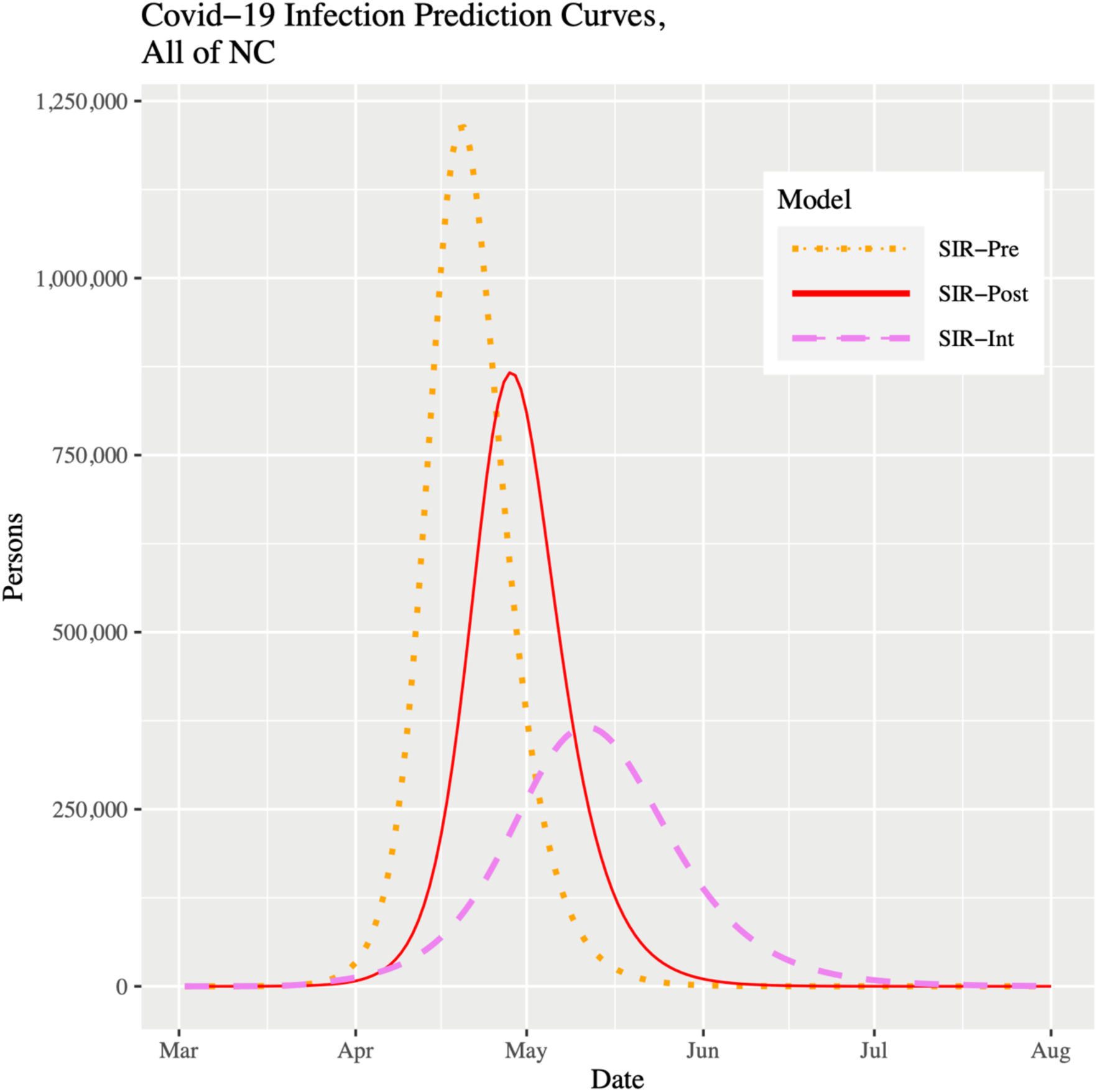
Infection prevalence prediction curves for NC up to August 1, 2020.

**Figure 8:**
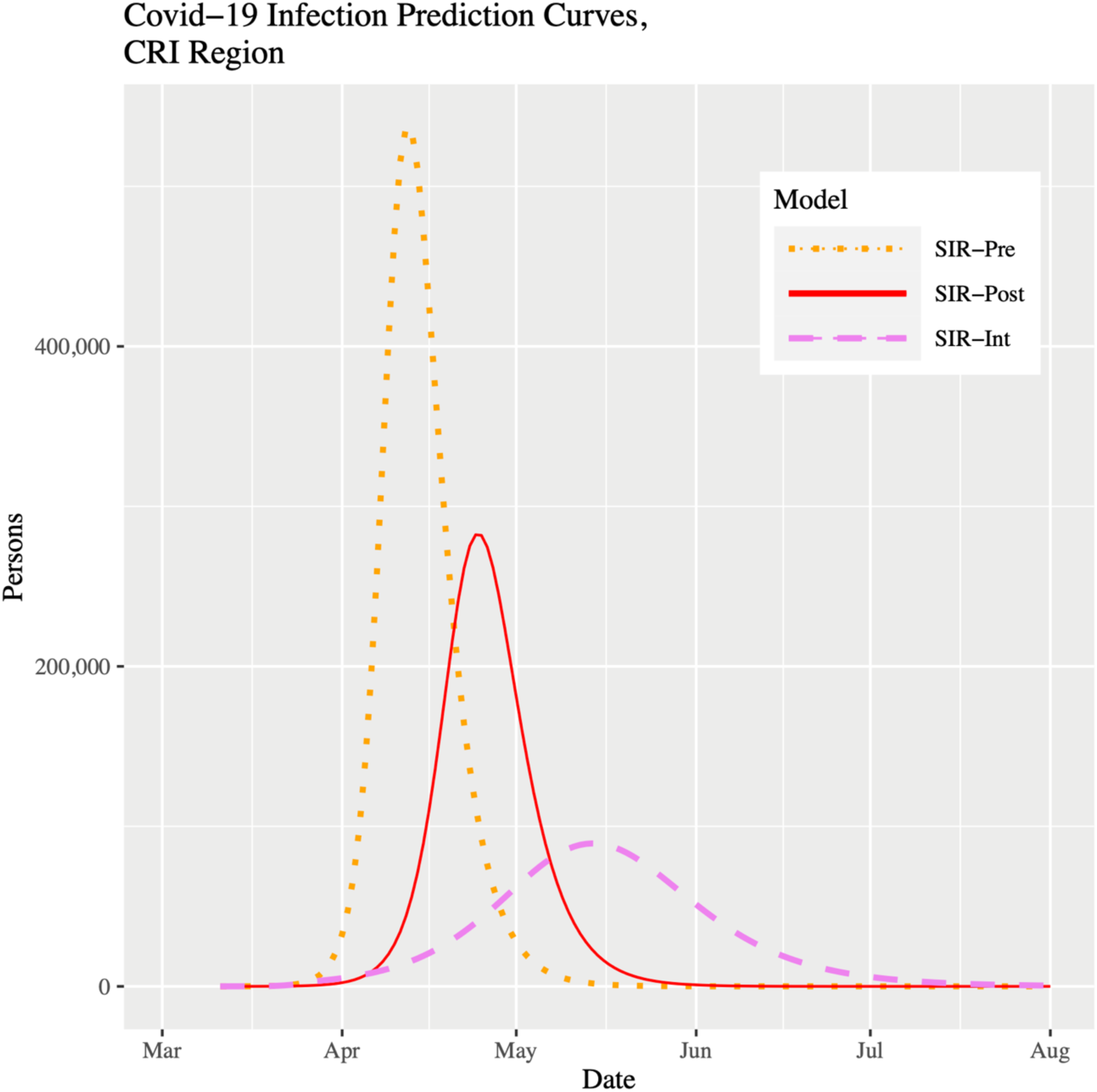
Infection prevalence prediction curves for the CRI up to August 1, 2020.

**Table 3:** Summary table describing infection under three different models in NC and the CRI.

| Peak Kinetics |  |  |  |  |  |  |  |
| --- | --- | --- | --- | --- | --- | --- | --- |
| Location | Model | 2020 Date | $\widehat{S}$ | $\widehat{I}$ | $\widehat{R}$ | Peak % Infected | Final % Infected |
| NC | SIR-Pre | Apr 20 | 5,673,270 | 1,213,190 | 3,601,625 | 12% | 73% |
| NC | SIR-Post | Apr 28 | 6,614,437 | 866,404 | 3,007,244 | 8% | 65% |
| NC | SIR-Int | May 11 | 7,913,011 | 366,037 | 2,209,037 | 3% | 46% |
| CRI | SIR-Pre | Apr 12 | 1,142,320 | 537,031 | 864,690 | 21% | 87% |
| CRI | SIR-Post | Apr 24 | 1,488,530 | 282,257 | 773,254 | 11% | 72% |
| CRI | SIR-Int | May 14 | 1,911,343 | 89,324 | 543,374 | 4% | 46% |

Figures 9 and 10 show plots of the three fitted models’ removal curves for NC and the CRI, respectively, projected out to the beginning of August. These plots support what we have observed so far. With the continued intervention, the removal curves are beginning to collapse, which is a behavior we would expect. For the SIR-Int model, both locations show a removal plateau being reached roughly around the beginning of July.

**Figure 9:**
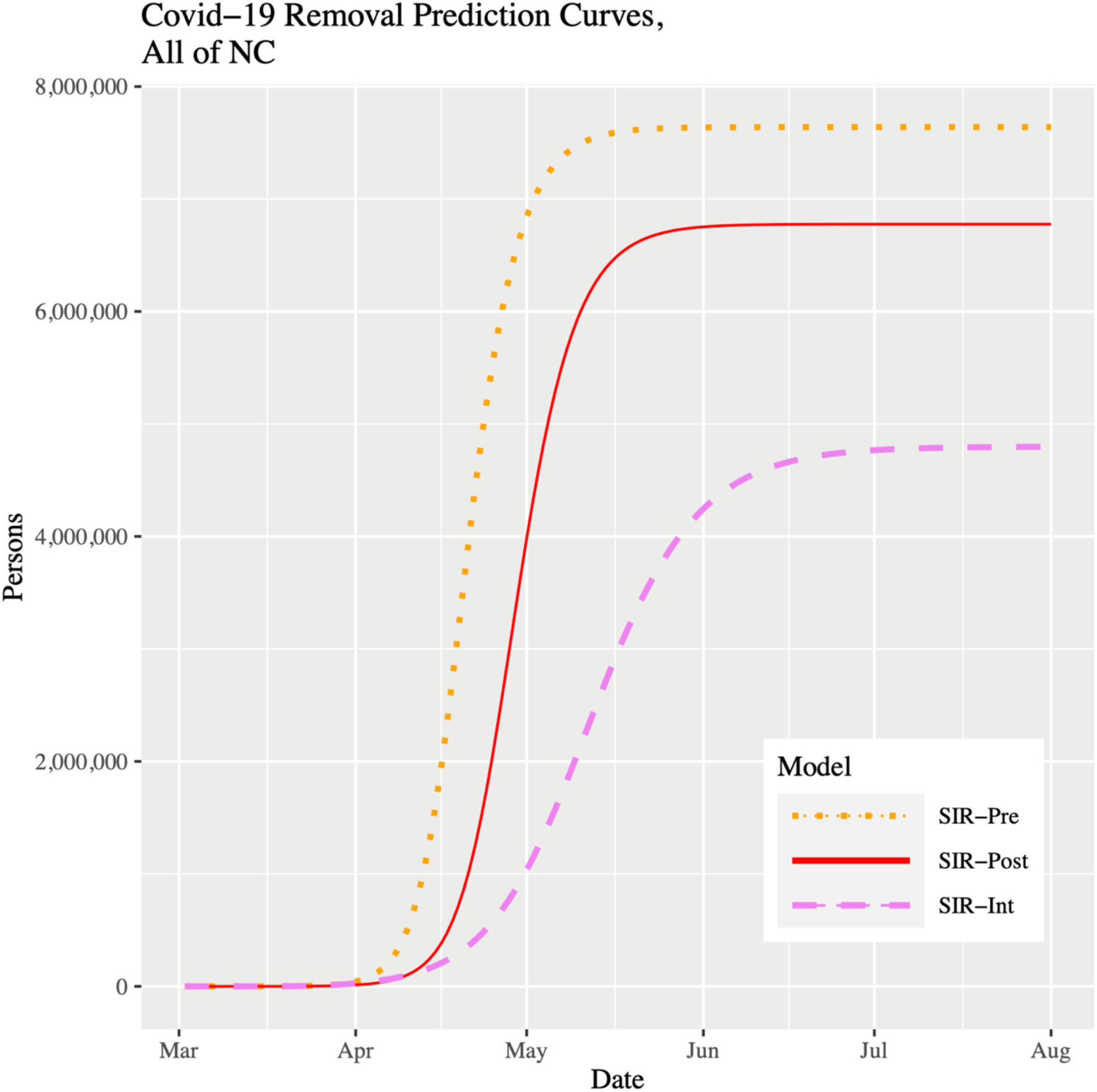
Removal prevalence prediction curves for NC up to August 1, 2020.

**Figure 10:**
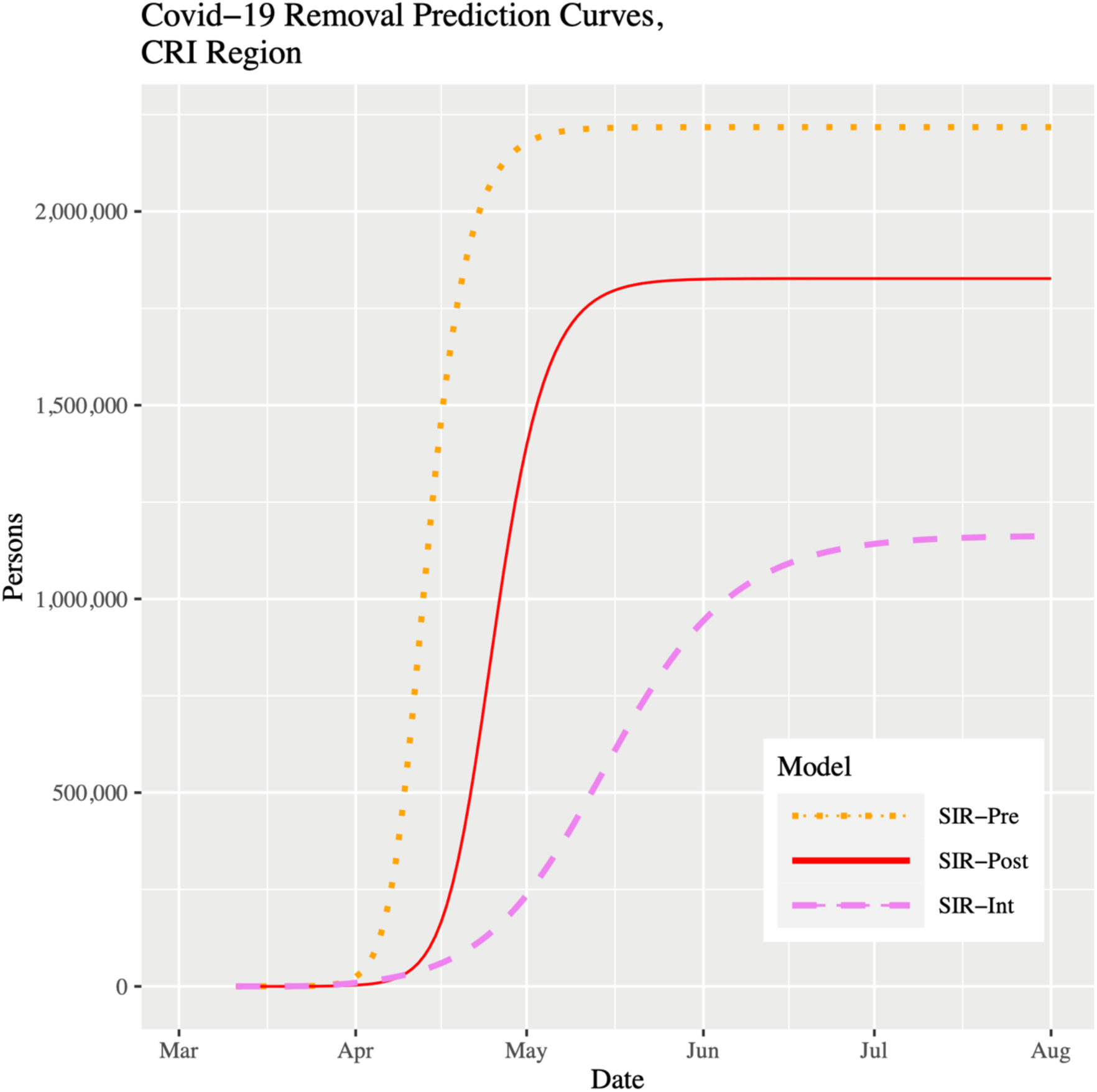
Removal prevalence prediction curves for the CRI up to August 1, 2020.

## Discussion

### Principal Results

In terms of model fitting, we state several observations. The SIR-Pre model represents a “worst case” scenario, as if the disease were allowed to run its course. Hence, early in a pandemic like this, it serves a useful purpose to help leaders understand the consequences of taking no action, or delayed action on implementing public health interventions. Beyond that, a basic SIR model, especially one that is used after being fit only to early pandemic data, imparts no further value for informing pandemic response planning, and indeed may provide errant forecasts. This diminished value also holds true when a basic SIR model is fit to contemporary data, yet ignores the effect of a public health intervention, as demonstrated by the SIR-Post model. Eventually, both such models will provide a poor fit to the data. Because the behavior of any epidemic is dynamic, any model requires constant monitoring, assessment of fit to local data, and evaluation of efficacy as new data are collected or additional research becomes available. Our SIR-Int model provides an example where this attention to model fit and incorporation of regional influences allows for appropriate model adaption and careful calibration, thus generating the most accurate predictions available to guide regional decision making at the time.

Summarizing the effect of the intervention, the doubling time for both locations is substantially slower after the intervention, with the CRI doubling time estimate (4.70 days) now being greater than for NC (4.01 days). The stay-at-home orders strongly appear to be working as intended as the infection curves for both locations are now becoming flatter (and <u>shrinking</u>), with peak infection prevalence now being pushed towards mid-May, both location’s recovery curves starting to fall, and measurable intervention effects on the hazard ratio and *R*_0_. It is interesting to note that our results match rigorous Monte Carlo simulation studies we conducted weeks beforehand.

If we compare the two locations, the estimated *R*_0_ of 2.36 for the CRI prior to March 26 is more typical of the range of *R*_0_ values in the literature for COVID-19, while the value of 1.79 for NC is substantially lower. This could be attributed to the fact that the CRI contains the largest city in NC, and one of the US’s busiest airports, setting the stage for this region to have become another COVID-19 hotspot. It is interesting to note that the NC SIR-Int model showed a better fit when the changepoint was also set to March 26, rather than March 30 when the statewide stay-at-home order went into place. One possible explanation for this could be that as the pandemic began in earnest, the general population’s fear of the virus also increased, perhaps causing most NC citizens to shelter-in-place prior to the order going into effect. Another explanation is that Mecklenburg County accounts for almost 11% of the NC population and so the effect of the county order directly impacted adjoining counties in the CRI, thus influencing the observed effect at the state level. Two additional interesting observations highlight the critical influence of spatial variation. First, the CRI infection curve evidences relatively more flattening and a later peak infection date. Second, the intervention effect in the CRI also appears stronger. The likely explanations for these differences are the Mecklenburg County stay-at-home policy going into effect five days before the state order, the different reaction of the local population to the order and its related messaging, and innumerable other unknown covariates such as early canceling of religious services, public gathering policies, and canceling of elective medical visits and procedures.

### Limitations

There are limitations to the SIR model. Some take issue with its deterministic form, although one could fit a Bayesian SIR model to make it stochastic. Perhaps the biggest limitation is that *β* and γ could be time-varying due to different forms of intervention (enhanced personal protective measures and social distancing). However, as we have shown here, we can easily leverage pre-existing R functions to incorporate a changepoint that modifies the probability of transmission to acknowledge an important public health intervention. It is also possible to customize the SIR model within R to define more advanced and different transition processes, and then parameterize and simulate those models to accommodate insights from additional research. In this way, one can also examine “what if” scenarios or assess model robustness through sensitivity analysis. The SIR model is simple to understand and easier to fit, as opposed to other deterministic compartmental models (e.g., SEIR) or stochastic individual contact models [31]. However, these more advanced models will play an increasingly important role in forecasting and understanding the dynamics of this evolving pandemic.

The lack of widespread COVID-19 testing, both for symptomatic and asymptomatic individuals, presents a major limitation of unknown scale and implications to forecasting models [32, 33]. Data sources are known to undercount cases, only including asymptomatic illness by chance, and to define cases inconsistently, based on variable testing criteria, between and within geographies. Collectively, these contribute to imperfect detection. As a result, high-level models may not comprehend the full extent of the outbreak, creating challenges in producing accurate forecasts. Our decision to base our modeling strategy on estimated latent prevalence addresses this inconsistency by adjusting observed prevalence counts. Modeling only the observed prevalence has the effect of shifting the SIR curves ahead in time by several days or more. While our estimate of the detection probability (0.14) is heuristically motivated, a thorough search of the literature supports our use of this estimate as reasonable. Future work will focus on refining this estimate as new research appears and to allow it to vary as a function of time.

### Comparison with Prior Work

While there is a plethora of models that estimate the impact of COVID-19 in the US, there are far fewer that give localized projections. We note that our mid-May date for the peak infection curve is roughly 3-to-4 weeks later than the projection from the often-cited model from The Institute for Health Metrics and Evaluation [34]. The later uses a Bayesian generalized nonlinear mixed model to examine cumulative death rates and assumes a strict social distancing policy is in place. Using data up until March 13, Columbia University reported a mid-May peak time for NC under no control measures and a start of July peak time under some control measures [35]. The authors caution that their metapopulation SEIR model is designed to capture national trends, and local projections should be viewed as broad estimates. Other models, such as the CHIME model from the University of Pennsylvania Health System, relied on data from three Pennsylvania hospitals to estimate hospital capacity and clinical demand and was not designed to capture changing regional mitigation strategies [36].

### Policy and Practice Implications

In the context of limited national policy guidelines to reduce COVID-19 transmission, provide resources for healthcare system pandemic preparedness and mitigate health consequences, state and local authorities must have reliable, timely, and geographically specific models to manage the unfolding crisis. We provided our local forecasts to health system leaders and public health officials to help guide regional planning. Because we regularly refit our models to local data, these served as a flexible tool enabling first proactive preparedness based on the initial pandemic trajectory, followed by timely pivoting of capacity planning to match the observed disease deceleration. Furthermore, locally accurate forecasts enhance the relevance of forecasting’s role in public health communication [37]. For example, the potential disease impact on local health system capacity may help communities understand the rationale for public health interventions; whereas, the positive effects of community mitigation may provide reinforcement for maintaining strategies like social distancing and enhanced hygiene.

Using regional and state data, we demonstrate how epidemiological modeling based on local context is critically important to informing pandemic preparedness for health systems and policy leaders. The results highlight the importance for such models to be created using local data, as opposed to running a simulation which makes many assumptions about the truth of parameter values. All models should be continuously re-calibrated, and adapted to the rapid, continuously changing situations inherent to a pandemic. A one size fits all approach to the underpinning forecasting model or reliance on data that does not incorporate local context, sets the stage for misguided forecasting. Additionally, our study shows that while a classic SIR model may perform well in the early days of the pandemic, it begins to lose relevance with the emergence of additional influences like social distancing and enhanced awareness of personal hygiene.

The SIR-Int model has high predictive accuracy based on data collected from March 2 to April 7 for both NC and the CRI and is able to demonstrate clear, compelling evidence of the efficacy of a stay-at-home order. By modeling estimated latent prevalence as we have done here, instead of observed prevalence, a lag delay in projecting peak infection can be avoided, reducing the consequences to leaders who require an accurate timeline for planning purposes (e.g., surge planning of hospital beds, supplies, and personnel).

## Conclusions

All other things being equal, if residents continue to observe the stay-at-home orders, maintaining attention to social distancing and increased personal hygiene, then this wave of the COVID-19 outbreak would essentially be over by mid-July. It is possible that we could see continued flattening and shrinking of the infection curve in which case our forecast results would adapt commensurately. It is also possible that infection prevalence could oscillate at a low level over time, in which case more advanced modeling and methods would be needed. Our results highlight the importance of incorporating local context into pandemic forecast modeling, as well as the need to remain vigilant and informed by the data as we enter into a critical period of the outbreak. While there will regrettably still be tragic loss of life and many NC citizens infected by coronavirus, this scenario pales in comparison to what could have been a far worse conclusion.

## Data Availability

Data used in this study were obtained from the North Carolina Department of Health and Human Services website, which can be accessed here: https://www.ncdhhs.gov/divisions/public-health/covid19/covid-19-nc-case-count.

## Acknowledgements

PT takes full responsibility for the integrity and accuracy of the statistical analysis and assembling the manuscript; SHC generated the map and was involved in discussions on statistical analysis; YT and MS drafted the abstract and discussion; MK and JP drafted the introduction; PT drafted the methods, results, and part of the discussion; MK, MS, YT and PT assembled references; YT, SHC, MS, and AW performed critical revision of the manuscript for important intellectual content; PP collected the data and drafted the bibliography; and AW supervised the study.

